# Altered reward network responses to social touch in major depression

**DOI:** 10.1101/2021.11.25.21266854

**Authors:** Clemens Mielacher, Dirk Scheele, Maximilian Kiebs, Laura Schmitt, Torge Dellert, Alexandra Philipsen, Claus Lamm, René Hurlemann

**Affiliations:** Research Section Medical Psychology, Department of Psychiatry and Psychotherapy, University Hospital Bonn, Bonn, Germany; Department of Psychiatry, School of Medicine & Health Sciences, University of Oldenburg, Bad Zwischenahn, Germany; Institute of Medical Psychology and Systems Neuroscience, University of Münster, Münster, Germany; Department of Psychiatry and Psychotherapy, University Hospital Bonn, Bonn, Germany; Social, Cognitive and Affective Neuroscience Unit, Department of Cognition, Emotion, and Methods in Psychology, Faculty of Psychology, University of Vienna, Vienna, Austria; Research Center Neurosensory Science, University of Oldenburg, Oldenburg Germany

**Author notes:** Corresponding author: Clemens Mielacher, Research Section Medical Psychology, Department of Psychiatry and Psychotherapy, University Hospital Bonn, Venusberg-Campus 1, 53127 Bonn, Germany, **Email:**.

**Keywords:** Major depressive disorder, social touch, reward, striatum, fMRI

## Abstract

**Introduction:** Affective touch is highly rewarding and an integral part of social relationships. Major depressive disorder (MDD) is characterized by severe impairments in reward processing, but the neural effects of social touch in MDD are still elusive.

**Objective:** We aimed to determine whether the neural processing of social touch is impaired in MDD and to assess the impact of antidepressant therapy.

**Methods:** Before and after antidepressant treatment, 53 MDD patients and 41 healthy controls underwent functional magnetic resonance imaging (fMRI) while receiving social touch. We compared neural responses to social touch in the reward network, behavioral ratings of touch comfort and general aversion to interpersonal touch in MDD patients to controls. Additionally, we examined the effect of treatment response on those measures.

**Results:** Clinical symptoms decreased after treatment and 43.4% of patients were classified as responders. Patients reported higher aversion to social touch and lower comfort ratings during the fMRI paradigm than controls. Patients showed reduced responses to social touch in the nucleus accumbens, caudate nucleus and putamen than controls, both before and after treatment. Non-responders exhibited blunted response in the caudate nucleus and the insula compared to responders, again irrespective of treatment.

**Conclusions:** These findings confirm our hypothesis that interpersonal touch as an indicator of social reward processing is impaired in MDD. Persistent dysfunctional processing of social touch despite clinical improvements may constitute a latent risk factor for social withdrawal and isolation. New treatment approaches are necessary to specifically target social reward processing and disturbed body awareness in MDD.

## Introduction

Major depressive disorder (MDD) is one of the most common mental disorders and a leading cause of years lived with disability [1]. A core symptom of MDD, according to both DSM-V and ICD-10 criteria, is anhedonia, an array of deficits impacting various hedonic functions such as desire, motivation and pleasure [2]. Patients suffering from anhedonia show overall poorer treatment response [3,4], possibly because established pharmacotherapies, particularly selective serotonin reuptake inhibitors (SSRIs), are not well suited to treat motivational and reward-related dysfunctions in depression [5,6]. On a neurobiological level, anhedonia has been associated with the reward network (for an overview, see ref. [7]). Meta-analytical evidence from neuroimaging studies shows that patients with MDD exhibit reduced responses to monetary incentives and happy faces in various reward network nodes, such as the nucleus accumbens, caudate, putamen, insula and orbitofrontal cortex [8–10]. Moreover, higher reward sensitivity is associated with better outcome after psychotherapeutic interventions. [11]

Social interactions are considered natural rewards [12] and activate the reward network in healthy participants [13–16]. Even though MDD patients often suffer from impairments in social functioning (for an overview, see [17]), few studies have probed the processing of social reward in MDD [18,19]. For instance, social touch can be inherently rewarding and is an integral part of nonverbal social communication and bonding [20,21], but it is still elusive whether MDD also modulates the processing of rewarding interpersonal, tactile stimulation.

Social distancing measures in the era of COVID-19 have vividly demonstrated the importance of interpersonal touch and the consequences of its absence. Social touch deprivation during the pandemic has been linked to increased anxiety and loneliness and resulted in a craving for interpersonal touch [22,23]. The processing of touch is mediated by different pathways in the nervous system. Myelinated Aβ-fibers enable rapid central processing and convey discriminative information, allowing for prompt responses to a stimulus. These fibers are activated by fast tactile stimulation, whereas unmyelinated C-tactile (CT) afferents respond to slow, caressing stimulation that corresponds to rewarding and affective properties of touch [24]. Social touch is intrinsically rewarding [25,26] and its rewarding nature seems to be mediated by the endogenous opioid system, in particular by the µ-opioid receptor system, which is involved in both non-social [27,28] and social reward [18,29–31]. In healthy participants, social touch increases µ-opioid receptor activity in the striatum and the insula among other regions [32], and opioid receptor antagonists modulate the motivation to obtain social touch [33] and its perceived pleasantness [34]. Functional magnetic resonance imaging (fMRI) studies provide further evidence for the effect of interpersonal touch on the reward circuit. Being touched by another person, but not self-produced touch, increases neural activation in the caudate nucleus [35]. Intranasal oxytocin, a neuropeptide crucially involved in social bonding, increases nucleus accumbens activity when participants believe they are being touched by their romantic partner [36]. Similarly, increased pleasantness ratings and striatal activity have been observed when heterosexual male participants believe social touch is being delivered by a female as opposed to a male experimenter [37,38]. Striatal response to affective touch seems to increase with age [39].

The rationale of the present study was to probe whether MDD is associated with impaired processing of social touch as an indicator of social reward. We therefore examined patients with MDD before and after a multi-week course of antidepressant treatment and compared them to healthy controls who were examined over the same period. We employed a social touch fMRI paradigm, during which participants rated the comfort of affective (i.e., slow) and discriminative (i.e., fast) touch. Additionally, we assessed depressive symptom severity over the course of the study in MDD patients. We expected MDD patients to perceive social touch as less comfortable and to display decreased neural responses to social touch in reward-associated regions compared to healthy controls, particularly in the nucleus accumbens, caudate nucleus, putamen and insula. We further hypothesized that these MDD-related impairments would decrease after treatment. Since anhedonia is associated with worse treatment outcome, we expected that non-responders to antidepressant therapy would report lower comfort ratings and exhibit lower neural responses to social touch compared to responders. We assumed that these effects would be particularly pronounced in response to slow affective as opposed to fast discriminative touch.

## Materials and Methods

### Participants and study design

Between June 2016 and April 2018, 53 patients with MDD (27 female, age 41.58 ± 13.09 years) and 41 healthy controls (22 female, age 40.61 ± 13.22 years) participated in this study (Table 1). To participate in this registered study (https://clinicaltrials.gov/show/NCT04081519), all patients had to meet DSM-IV criteria for unipolar MDD as diagnosed by an experienced psychiatrist and verified by the Mini-International Neuropsychiatric Interview (MINI) [40], and were in-patients at the Department of Psychiatry, University Hospital Bonn, Germany. Exclusion criteria for all participants were suicidal ideation, psychotic symptoms, bipolar depression, substance abuse, eating disorders, post-traumatic stress disorder, personality disorders, neurological disorders and MRI contraindications. For healthy controls, additional exclusion criteria were any lifetime axis I or II psychiatric disorders and any past or current psychopharmacological medication. To assess a possible history of abuse and neglect, we administered the Childhood Trauma Questionnaire (CTQ) [41]. General attitude toward touch was assessed using a Social Touch Questionnaire (STQ) [42].

**Table 1.**
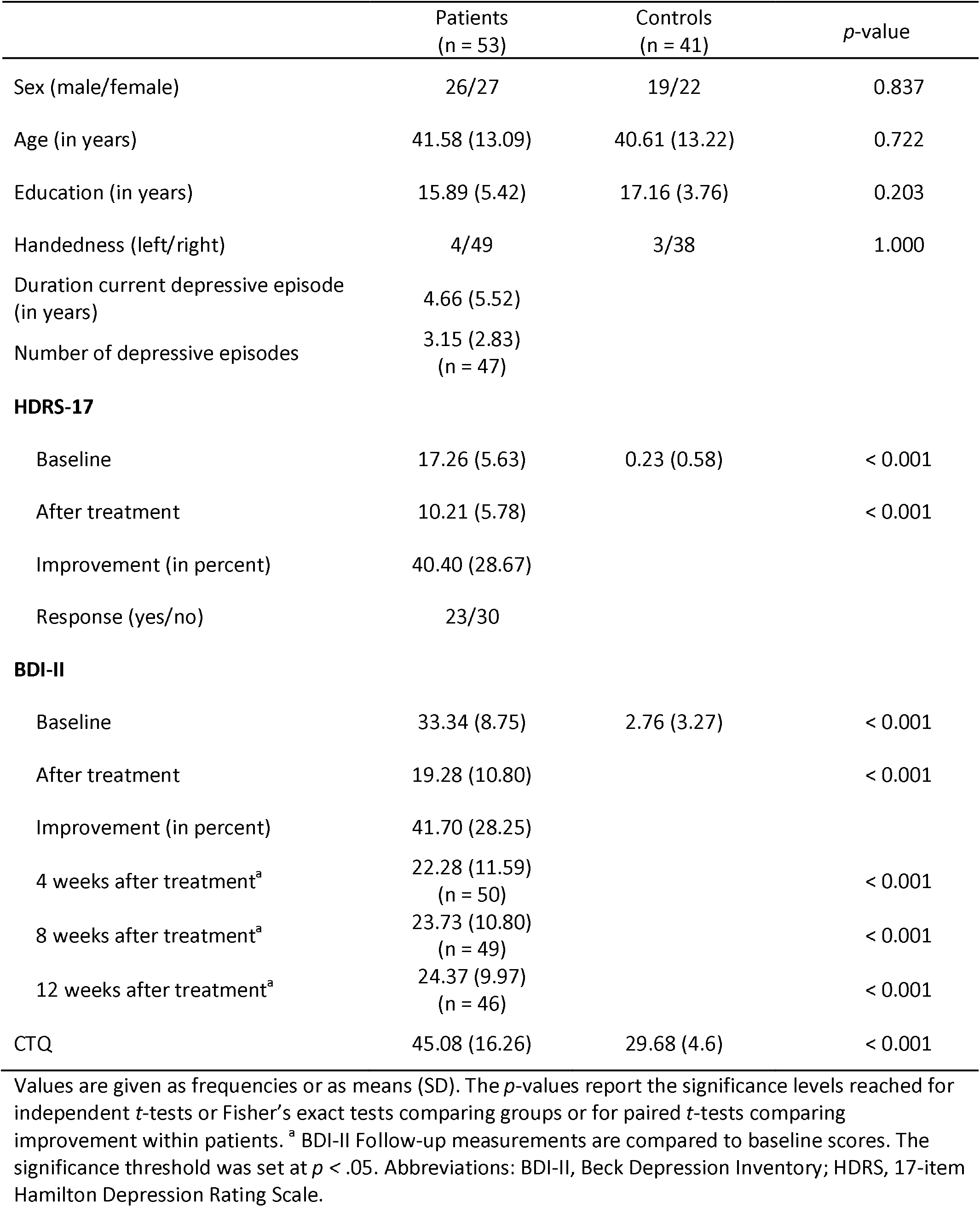
Demographic and clinical data for patients and controls.

Patients underwent MRI scanning within 1–3 days after admission to the clinic and, again, 24 days later; accordingly, controls were examined twice at the same interval. For the duration of the study, patients received treatment according to current guidelines for MDD [43] (cf. supplementary information). To quantify clinical improvement, trained raters assessed depressive symptom severity on a weekly basis using the 17-item Hamilton Depression Rating Scale (HDRS-17) [44]. As a measure of self-assessed depression severity, the Beck Depression Inventory (BDI-II) [45] was administered before and after the treatment course and every four weeks over a 12-week follow-up period.

### Social touch paradigm

For the fMRI scans, we employed an adapted version of an established paradigm [46,47], in which tactile stimulation was manually applied to participants at different speed levels. Stimulation was administered by an experimenter who performed vertical strokes with cotton gloved hands over 20-cm zones on the participants’ shins that were marked prior to the fMRI scan. During the 4-s touch, the complete zone was covered either with a single stroke at a speed of 5 cm/s (slow, affective touch) or with four repeated strokes at a speed of 20 cm/s (fast, discriminative touch). Slow is experienced as more pleasant than fast touch [48] and specifically elicits responses by CT afferents, which are associated with rewarding properties of touch [24]. The experimenter was trained to keep stimulation pressure constant at both speed levels and received audio cues via headphones during the experiment to ensure constant stimulation velocity. No stimulation occurred during the no touch control condition. Each condition was repeated 20 times in randomized order. Each trial was initiated with the presentation of a white fixation cross (3 s). Fast and slow touch trials were then announced by the color of the fixation cross changing to blue (1 s). After each trial, the participant rated the comfort of the tactile stimulation on a 100-point visual analogue scale that ranged from not at all comfortable (0) to very comfortable (100) and was presented for a maximum of 5 s. To minimize context effects, participants were not informed about the identity of the person administering the stimulation and the opening of the scanner was covered with a blanket during the experiment.

### MRI data acquisition

Functional and structural MRI data were acquired on a 1.5 T Siemens Avanto MRI system (Siemens, Erlangen, Germany) equipped with a 12-channel standard head coil at the Life & Brain Centre, Bonn, Germany. T2*-weighted gradient-echo planar images (EPI) images with blood-oxygen-level-dependent (BOLD) contrast were acquired during the social touch task (voxel size = 3×3×3 mm; TR = 3000 ms; TE = 50 ms; flip angle = 90°; FoV = 192 mm, matrix size = 64×64; 35 axial slices; ascending slice order with interslice gap of 0.3 mm). The first five volumes of each functional time series were discarded to allow for T1 equilibration. Additionally, a field map (voxel size = 3×3×3 mm; TR = 460 ms; TE_fast_ = 4.76 ms; TE_slow_ = 9.52 ms; flip angle = 60°; matrix size = 64×64; 35 axial slices; interslice gap of 0.3 mm) was acquired to correct for inhomogeneities of the magnetic field during preprocessing. Subsequently, a high-resolution structural image was acquired using a T1-weighted 3D MRI sequence (voxel size = 1×1×1 mm; TR = 1660 ms; TE = 3.09 ms; flip angle = 15°; FoV = 256 mm; matrix size = 256×256, 160 sagittal slices).

### Data analysis

Data analyses focused on the comparison of patients with healthy controls and on differences between responders and non-responders to antidepressant treatment. The criterion for clinical response was defined as a ≥ 50% reduction in HDRS-17 scores.

The fMRI data were preprocessed and analyzed using SPM12 software (Wellcome Trust Center for Neuroimaging, London, UK; http://www.fil.ion.ucl.ac.uk/spm) running in MATLAB R2010b (The MathWorks, Natick, MA). The functional data were realigned, initially to the first image in the time series, then to the mean of all images, and unwarped using the field map data. They were then coregistered to the anatomical volume acquired pre-treatment and normalized based on probabilistic tissue segmentation into 2-mm stereotaxic Montreal Neurological Institute (MNI) space. Subsequently, the images were smoothed using a 4-mm full width at half maximum (FWHM) Gaussian kernel. Two patients and one control had to be excluded from further fMRI analysis due to excessive head movement (> 3 mm or °) during data acquisition. This resulted in a sample size of 51 patients and 40 controls. A two-level random effects approach based on the general linear model as implemented in SPM12 was used for statistical analysis. After preprocessing, conditions based on combinations of stimulus (fast touch, slow touch) and time (pre-treatment, post-treatment) were entered into a GLM for each participant together with a constant term and six realignment parameters per session to account for subject motion. On the first level, we subtracted the respective no touch control regressor from the experimental regressors for each participant and condition. On the second level, we conducted two separate analyses of variance (ANOVA) to compare patients with controls, and responders with non-responders. For each analysis, we entered the first level contrasts in separate flexible factorial models to compute the within-subject main effects of speed (fast touch, slow touch) and time (pre-treatment, post-treatment), the between-subjects main effects of group (patients, controls) or response (responders, non-responders), and their respective interactions. For each analysis, we used multiple models to partition variance in SPM as recommended when using group-level repeated measurement designs [49].

To validate the effect of the social touch paradigm, we performed a whole-brain analysis of the control group with an initial height threshold of *p <* .001. Peak-level *p*-values were then family-wise error (FWE) corrected for multiple comparisons and *p <* .05 was considered significant.

The main analysis focused on a set of bilateral a priori defined regions of interest consisting of the nucleus accumbens, caudate nucleus, putamen and anterior and posterior insula. These regions were defined based on the automated anatomical labelling atlas 3 [50]. The peak-level threshold for significance was set to *p <* .05, FWE-corrected for multiple comparisons based on the size of each region of interest.

Behavioral data were analyzed using SPSS Statistics Version 27 (IBM Corp., Armonk, NY, USA) and all tests were two-tailed. To test for clinical improvement, a repeated measures ANOVA was performed for HDRS-17 ratings. In line with the fMRI analyses, we conducted separate mixed-design ANOVAs of social touch comfort ratings with touch speed (slow, fast) and time (pre-treatment, post-treatment) as within-subject factors and either group (patients, controls) or response (responders, non-responders) as a between-subjects factor to compare patients with controls or responders with non-responders, respectively. The threshold for significance was set to *p <* .05, and *p*-values were Bonferroni-adjusted if appropriate (*P*_*corr*_). Greenhouse-Geisser correction was applied in cases of lack of sphericity. A moderation analysis was conducted to examine the effect of potential confounders (age, sex, CTQ scores) on our analyses (cf. supplementary information). Partial eta-squared and Cohen’s *d* were calculated as measures of effect size. Pearson’s product-moment correlation was used to test associations between fMRI peak-voxel parameter estimates from the region of interest analysis and comfort ratings, social touch aversion, HDRS-17 baseline scores and HDRS-17 item number 7 as a measure of baseline anhedonia. This item assesses “loss of interest in activities”, “decrease in actual time spent on activities” and “experiencing pleasure” [44].

## Results

### Behavioral results

Analysis of HDRS-17 scores (shown in Fig. 1) showed a significant reduction over time (*F*_(2.59, 134.89)_ = 36.82, *p <* .001, η_p_^2^ = 0.42) in patients, 23 (43.4%) of whom met the criterion for a clinical response.

**Fig. 1.**
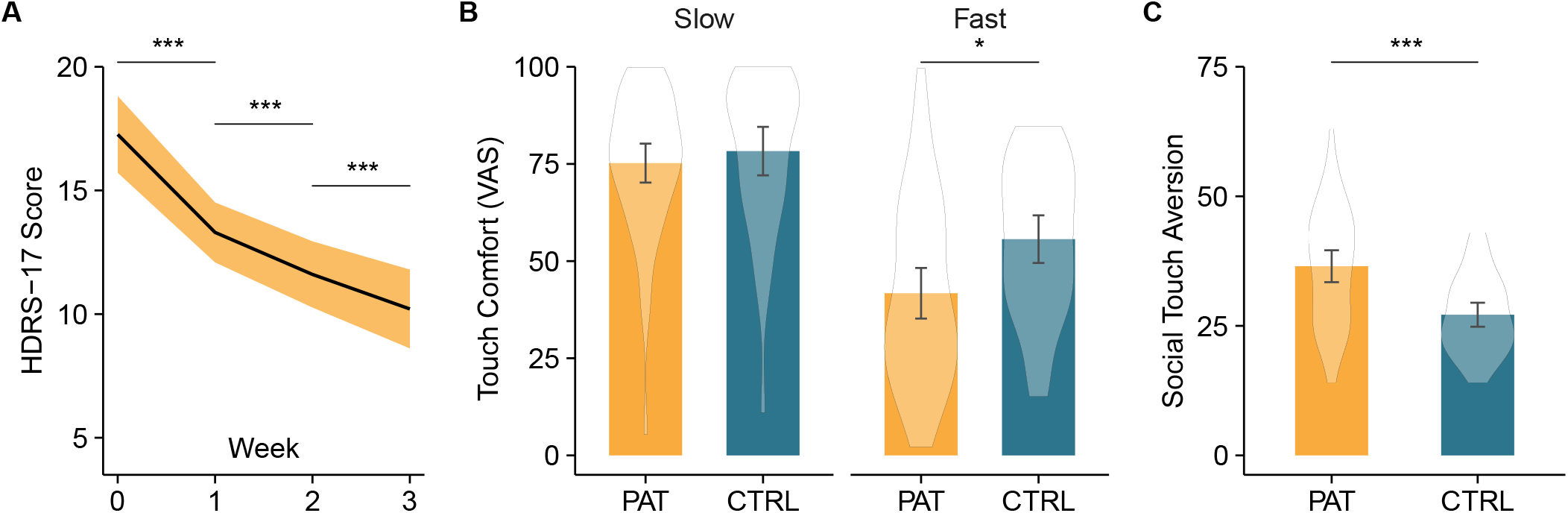
Depression symptom severity as measured by Hamilton Depression Rating Scale (HDRS-17) scores decreased over the treatment course (**A**). Patients rated fast but not slow touch as significantly less comfortable than controls (**B**). At baseline patients reported a higher aversion to social touch than controls (**C**). Indicated *p*-values are Bonferroni corrected. Violin plots are kernel density plots comparable to histograms with infinitely small bin sizes. The ribbon and error bars indicate 95%-confidence intervals. Abbreviations: CTRL, controls; PAT, patients; VAS, visual analogue scale. \**p <* .05, \*\*\**p <* .001.

Analysis of social touch comfort ratings revealed main effects of speed (*F*_(1,92)_ = 99.46, *p <* .001, η_p_^2^ = 0.52) and group (*F*_(1,92)_= 7.12, *p =* .009, η_p_^2^ = 0.07, shown in Fig. 1). As expected, comfort ratings were higher after slow, affective touch than after fast, discriminative touch. Patients overall rated social touch as less comfortable than control participants. A trend toward an interaction between group and speed (*F*_(1,92)_ = 3.70, *p =* .058, η_p_ ^2^ = 0.04) indicated that this difference was only present after fast (*t* = 3.06, *p*_*corr*_ *=* .012, *d* = 0.64) but not slow touch (*t*_(92)_ = 0.79, *p*_*corr*_ > .999, *d* = 0.16). The analysis comparing responders and non-responders also revealed a significant main effect of speed (*F*_(1, 51)_ = 70.86, *p <* .001, η_p_^2^ = 0.58) with higher comfort ratings for slow touch, but no other significant main effects or interactions.

Patients reported a higher aversion to social touch as measured by STQ scores than controls (*t*_(89.88)_ = 4.89, *p <* .001, *d* = 0.97), while no difference was found between responders and non-responders (*t*_(51)_ = 0.08, *p* = .936, *d* = 0.02).

### fMRI results

In the control group, social touch relative to the no touch control condition revealed widespread activations in touch-processing networks at the whole-brain level including the insula, somatosensory cortex and supramarginal gyrus [51] (cf. supplementary information, Table S1).

In the region of interest analysis, patients showed diminished neural response to interpersonal touch irrespective of touch velocity and time (pre vs. post treatment) in the bilateral nucleus accumbens (peak MNI coordinates (x, y, z): -6, 16, -4; *F*_(1, 89)_ = 15.59, *p*_*FWE*_ = .010, η_p_^2^ = 0.14; MNI: 4, 14, -2; *F*_(1, 89)_ = 11.68, *p*_*FWE*_ = .041, η_p_^2^ = 0.11; shown in Fig. 2A) and in the bilateral caudate nucleus (MNI: -14, 20, 12; *F*_(1, 89)_ = 21.88, *p*_*FWE*_ = .005, η_p_^2^ = 0.19; MNI: 10, 10, 14; *F*_(1, 89)_ = 21.64, *p*_*FWE*_ = .006, η_p_^2^ = 0.20; shown in Fig. 2B) compared to controls. Reduced activity in both regions was associated with a general aversion toward social touch as assessed by the STQ (left nucleus accumbens: *r*_(89)_ = -0.28, *p =* .008; right caudate nucleus: *r*_(89)_ = -0.25, *p =* .017). Responses in the bilateral caudate nucleus negatively correlated with baseline anhedonia in patients (left: *r*_(49)_ = -0.44, *p =* .001; right: *r*_(49)_ = -0.32, *p =* .021). Since none of the correlations survived Bonferroni correction, the results should be interpreted with caution. Furthermore, we found a significant interaction between speed, time and group in the left putamen (MNI: -28, 0, 2; *F*_(1, 89)_ = 19.23, *p*_*FWE*_ = .016, η_p_^2^ = 0.18). Post-hoc tests revealed decreased responses to fast touch in patients compared to controls at baseline (*t*_(89)_ = 3.06, *p*_*corr*_ *=* .036, *d* = 0.65) but not after treatment (*t*_(89)_ = 0.38, *p*_*corr*_ > .999, *d* = 0.08).

**Fig. 2.**
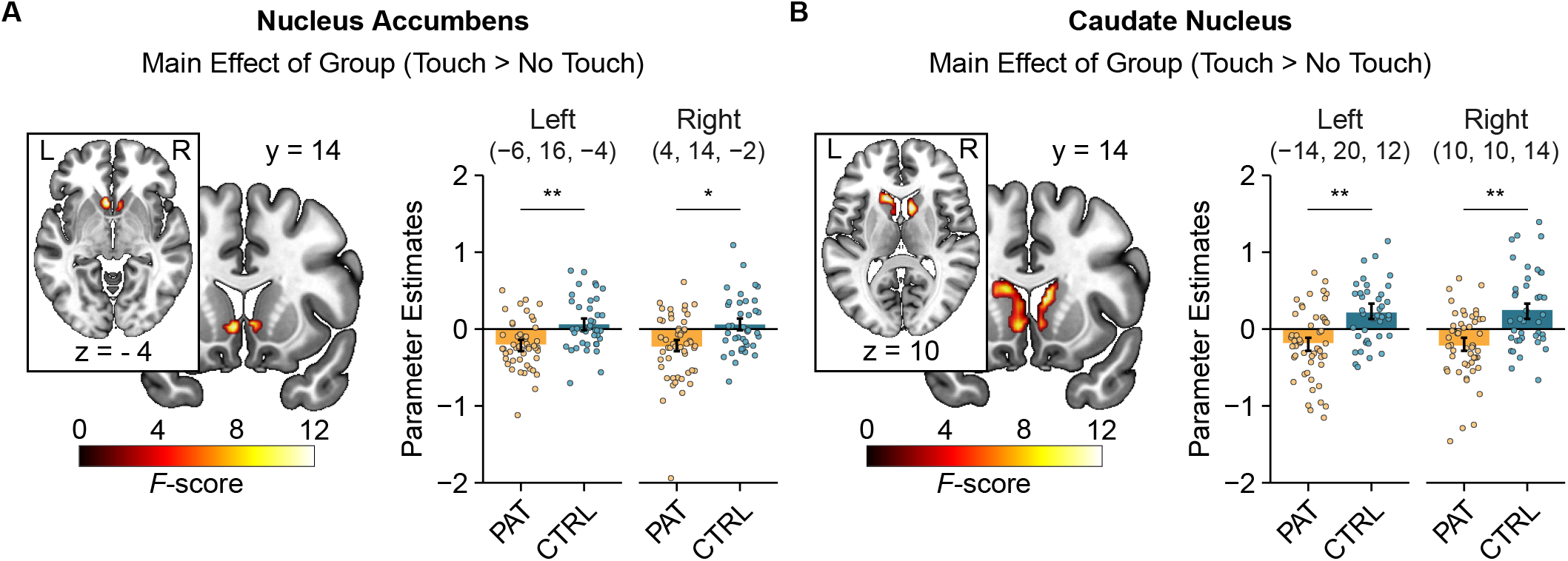
Patients exhibited decreased neural responses to social touch in the bilateral nucleus accumbens (**A**) and caudate nucleus (**B**) across time (i.e. before and after treatment) compared with healthy controls. Significant clusters are displayed at a peak-level threshold of *p <* .05 uncorrected. Parameter estimates are displayed for peak voxels. Error bars indicate 95%-confidence intervals. Abbreviations: CTRL, controls; PAT, patients. \**p <* .05, \*\**p <* .01.

Secondly, we examined the effect of treatment response. The main effect of treatment response indicated reduced activity during social touch in the right caudate nucleus (MNI: 22, 20, 12; *F*_(1, 49)_ = 17.86, *p*_FWE_ = .039, η_p_ ^2^ = 0.26, shown in Fig. 3A) in non-responders compared to responders. A significant interaction between speed and group in the left anterior insula (MNI: -26, 26, 2; *F*_(1, 49)_ = 20.01, *p*_*FWE*_ =.022, η_p_^2^ = 0.30, shown in Fig. 3B) showed that non-responders exhibited reduced activation during slow touch compared to responders (*t*_(49)_ = 3.75, *p*_*corr*_ = .002, *d* = 1.06), but not during fast touch (*t*_(49)_ = 0.01, *p*_*corr*_ > .999, *d* < 0.01). For the interaction of speed, time and group, we found two significant clusters in the right putamen (MNI: 32, -2, -8; *F*_(1,49)_ = 19.33, *p*_FWE_= .032, η_p_^2^ = 0.28; MNI: 30, -6, 10; *F*_(1,49)_ = 18.20, *p*_FWE_ = .046, η_p_^2^ = 0.27). Post-hoc tests revealed no significant effects after Bonferroni correction (all *p*_*corr*_ > 0.05). See supplementary information for main effects of time and speed.

**Fig. 3.**
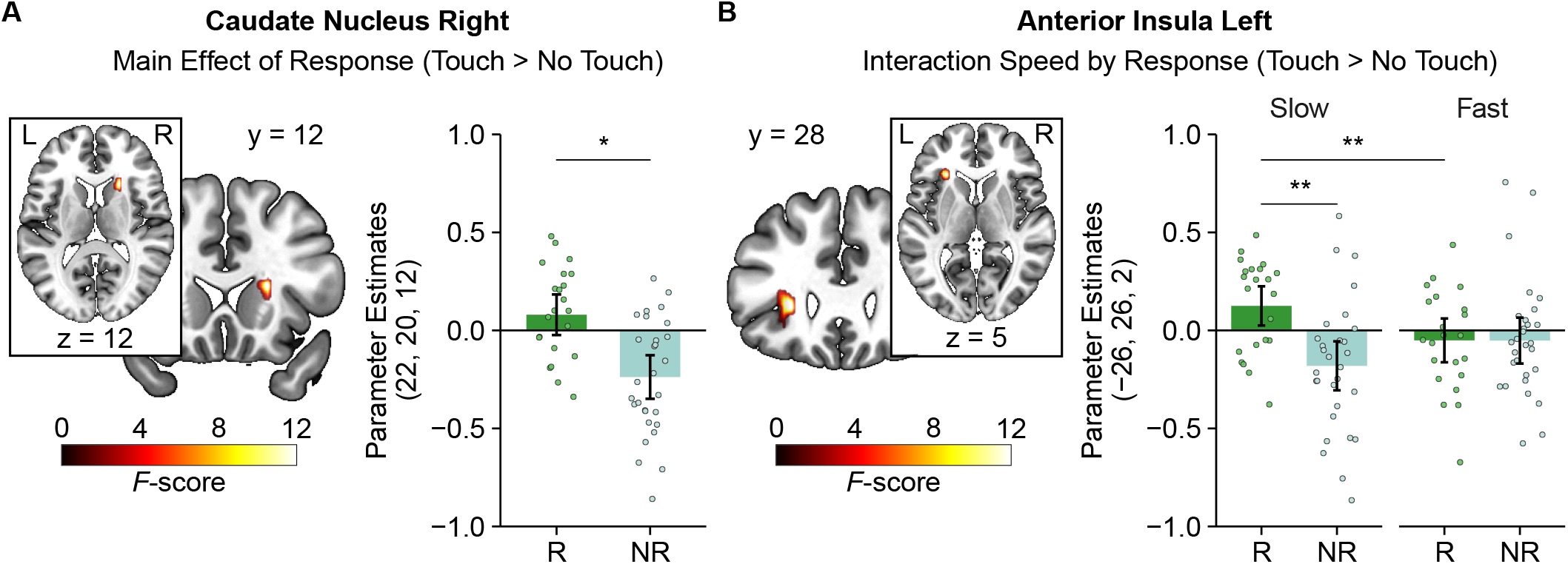
Treatment responders exhibited heightened neural responses to social touch in the right caudate nucleus across time compared with non-responders (**A**). Responses to slow touch in the left anterior insula were increased in responders across time compared with non-responders (**B**). Significant clusters are displayed at a peak-level threshold of *p <* .05 uncorrected. Parameter estimates are displayed for peak voxels. Indicated *p*-values are Bonferroni corrected. Error bars indicate 95%-confidence intervals. Abbreviations: NR, non-responders; R, responders. \**p <* .05, \*\**p <* .01.

The observed behavioral and neural effects of group were not significantly moderated by age or sex. We only found a significant suppressor effect of CTQ scores for the group effect on nucleus accumbens responses to social touch (cf. Supplement).

## Discussion/Conclusion

In the present study, we examined the processing of social touch as an indicator of social reward in depression. Confirming our first hypothesis, MDD patients reported a higher aversion to interpersonal touch, experienced it as less comfortable and exhibited reduced neural activation in the reward network compared to healthy controls. Specifically, we found decreased responses to social touch in the nucleus accumbens, caudate nucleus and putamen. Contrary to our expectations, the differences in the nucleus accumbens and caudate nucleus persisted even after treatment. In line with our second hypothesis, non-responders to antidepressant treatment displayed reduced activation in the caudate nucleus, anterior insula and putamen.

Unexpectedly, patients reported decreased comfort ratings compared to controls only after fast touch. This is in line with a study examining the effects of childhood maltreatment on the processing of social touch [46] and could be related to the specific rating of comfort instead of pleasantness [52]. Neural effects in the nucleus accumbens and caudate nucleus were independent of touch velocity, indicating that MDD-related impairments in reward-associated brain structures are not restricted to social touch with C tactile-optimized velocity. However, in line with our hypothesis, non-responders exhibited reduced reactivity in the insula specifically during slow touch compared to responders.

These findings corroborate the assumption that the processing of social reward in general [18,19,53] and of interpersonal touch in particular is impaired in MDD patients. The reported aversion to social touch in everyday life could indicate that diminished reward-associated responses to social touch could contribute to the emergence and reinforcement of social isolation in depression. MDD patients typically withdraw from their social circles, thus leading to smaller social network size [54,55] and increased loneliness [56,57], which is associated with more severe symptoms and a worse prognosis [58,59]. This disruption of social functioning can have devastating consequences, as both social isolation and loss of social support have been linked to suicidal outcomes [60,61].

Evidently, interpersonal touch is a crucial component of romantic relationships [62]. Impaired touch-associated reward might blunt the drive to seek physical closeness or even result in an avoidance of interpersonal touch, which could negatively affect sexuality and the overall satisfaction in romantic relationships [63–65]. Eventually, this might lead to separation, which is again a predictor for worse illness trajectories [66,67] and increased risk for suicidal behaviors [60].

Notably, the observed alterations of activity in the nucleus accumbens and caudate nucleus did not change over the treatment course. This could suggest a stable, phenotypical trait characterizing MDD patients that persists even after clinical improvement. This is in line with observations in remitted MDD patients who exhibit lasting impairments both in behavioral [68,69] and neural markers of reward processing [70–72], consistent with the persistence of anhedonia even after recovery from depression [73,74]. Correspondingly, we found a positive association between the response to social touch in the bilateral caudate nucleus and a marker for anhedonia in our data. Another explanation for the persistence of the impairments might be the relatively short time between the two fMRI sessions. While depressive symptoms went down by 40.4% across participants, a longer observation period perhaps would have allowed for further clinical improvement and behavioral adaptations. Likewise, a more pronounced impairment for slow touch was only evident in non-responders to treatment.

Considering the effect of response, we found reduced caudate nucleus and insula activation during social touch in non-responders both before and after treatment, indicating that those who show stronger impairments in striatal and insular reward processing might be less responsive to established antidepressant treatment, both in terms of clinical recovery and normalization of dysfunctional processing of social rewards. In the light of the devastating consequences that can arise from social isolation, this emphasizes the need for targeted interventions that focus on reward processing deficits. For instance, behavioral activation therapy [75] has been shown to be effective in the treatment of depression [76] and seems to affect striatal responses [77]. Furthermore, body-based interventions in the form of massage therapy [78] and body psychotherapy [79] are promising approaches to specifically target disturbed body awareness and desynchronization in depression [80,81].

Our findings should be interpreted in light of some limitations. Reward network activation during touch is in line with studies in healthy controls using various kinds of social touch conditions [32,35,37,38], but future studies are warranted to explore whether the observed impairments in MDD are specific to social touch or extend to the processing of non-social tactile stimulation. In addition, antidepressant treatment in this study was naturalistic and heterogeneous, and its particular influence on our findings therefore remains uncertain. However, the treatment was in line with current guidelines for the therapy of depression reflecting clinical realities.

In conclusion, our findings elucidate the role of social reward processing in depression and may help to refine the understanding of anhedonia, a core symptom of depression. Collectively, our results demonstrate an impairment of the experienced comfort of and neural response to social touch in patients with MDD. Moreover, these effects may constitute a risk factor for non-response and may persist even after recovery, leading to ongoing disruptions in social functioning. Future studies should corroborate these findings and might inform new treatment avenues targeting social reward and disturbances of body awareness.

## Data Availability

All data produced in the present study are available from the corresponding author C.M. upon reasonable request.

## Acknowledgements

The authors thank Paul Jung for his programming assistance as well as Anna Metzner, Lara Graute and Lea Köster for their help with data acquisition.

