## Supplement for "Altered reward network responses to social touch in major depression"

**Supplementary Methods**

*Subjects*

Patients between 18 and 60 years of age who fulfilled criteria for unipolar major depressive disorder for at least four weeks were eligible for inclusion. Physiological exclusion criteria were metal in the brain or the skull, a cardiac pacemaker or intracardiac lines, medication infusion devices, heart or brain surgery, pregnancy or any condition resulting in increased intracranial pressure, traumatic brain injury, a history of epilepsy, cerebral aneurysms, dementia, morbus Parkinson, Chorea Huntington, multiple sclerosis, stroke or transient ischemic attack (within the last 2 years). Psychiatric exclusion criteria included substance-induced depression, a history of substance abuse, psychotic episodes, bipolar disorder, anorexia, posttraumatic stress disorder (current or within the last 12 months), personality disorders, claustrophobia or previous antidepressant treatment with repetitive transcranial magnetic stimulation (rTMS), electroconvulsive therapy (within the last 3 months), vagus nerve stimulation or deep brain stimulation. All patients received concomitant multimodal treatment according to current MDD guidelines. The majority of patients (N = 47) received pharmacotherapy for the duration of the study: selective serotonin reuptake inhibitors (N = 18), selective serotonin-norepinephrine reuptake inhibitor (N = 15), atypical antidepressants (N = 32), atypical antipsychotics (N = 10), anticonvulsants (N = 11), tricyclic antidepressants (N = 5), levothyroxine (N = 4), antihistamines (N = 2), benzodiazepine (N = 1), lithium (N = 1), monoamine oxidase inhibitor (N = 1), norepinephrine reuptake inhibitor (N = 1). In addition, all patients underwent rTMS (for further information see [1]), group psychotherapy, and cognitive training [2].

See Table S1 for characterization of responders and non-responders.

*fMRI paradigm*

Stimulus presentation and response collection was implemented using Presentation 14 software (Neurobehavioral Systems, Albany, CA), liquid crystal display video goggles (Nordic NeuroLab, Bergen, Norway) and an MRI-compatible response box.

*fMRI whole-brain analysis*

To evaluate the effects of the touch paradigm, we performed a whole-brain analysis in controls using the first level contrasts [Touch > No Touch] and [No Touch > Touch] and one-sample *t*-tests on the second level. A threshold for significance of *P* < .05 was used, family-wise error corrected (FWE) for multiple comparisons. The results of this analysis can be found in Table S2.

*fMRI baseline analysis*

To corroborate our main findings, we conducted post-hoc analysis of baseline fMRI data. First level contrasts averaged over both speed levels at baseline were analyzed using independent t-tests comparing patients to controls ([patients > controls], [controls > patients]) and responders to non-responders ([responders > non-responders], [non-responders > responders]) using SPM. In accord with our main analysis, we focused on the same set of regions of interest. The peak-level threshold for significance was again set to *P <* .05, FWE-corrected for multiple comparisons based on the size of each region of interest.

*Statistical analysis*

Quantitative data were compared by repeated measures and mixed-design analyses of variance (ANOVA), and dependent and independent *t*-tests. Pearson's product-moment correlation was used for correlation analysis. Partial eta-squared was calculated as measures of effect size. For qualitative variables, Fisher’s exact tests were used. All reported *P*-values are two-tailed and values of *P* < 0.05 were considered significant.

*Moderation analysis*

We conducted a moderation analysis, using the PROCESS macro for SPSS, version 3.1 [3], to test for the potential confounding influence of age, sex and CTQ scores on the effect of group (patients, controls) and clinical response (responders, non-responders) on behavioral ratings and parameter estimates extracted from the fMRI analysis. All potential moderators were assessed individually in separate models. Moderation was assumed when the interaction between the predictor (group or response) and the moderator was significant. Additionally, Johnson-Neyman technique was applied to determine the conditional threshold of significance for any moderation effects.

**Supplementary Results**

*Clinical results*

When analyzing Hamilton Depression Rating Scale (HDRS) scores separately for responders and non-responders, both groups showed clinical improvement (responders: *F*_(2.11, 46.37)_ = 48.54, *P <* .001, η_p_^2^ = .69; non-responders: *F*_(2.13, 61.90)_ = 8.63, *P <* .001, η_p_^2^ = .23). Planned contrasts revealed continuous weekly improvement for responders (all *P*’s < .001), while non-responders only improved after the first week of treatment (*P =* .018) but not over the following weeks (all *P*’s > .200).

*fMRI results*

In addition to the effects reported in the main text, we found a main effect of time (pre vs. post treatment) in the right anterior insula, for the model comparing patients and controls. Activation to social touch decreased over the three weeks of treatment (peak Montreal Neurological Institute coordinates (x, y, z): 36, 26, −4; *F*_(1, 89)_ = 17.80, *P*_FWE_ = .024, η_p_^2^ = 0.17). We also found main effects of speed in the left nucleus accumbens (MNI: -6, 6, −4; *F*_(1, 89)_ = 12.97, *P*_FWE_ = .030, η_p_^2^ = 0.13) and the left posterior insula (MNI: -34, 2, 12; *F*_(1, 89)_ = 25.94, *P*_FWE_ = .001, η_p_^2^ = 0.22) both with heightened responses to slow touch compared with fast touch. Additionally, a significant main effect of speed in two clusters in the right posterior insula (MNI: 36, -14, 22; *F*_(1, 89)_ = 20.25, *P*_FWE_ = .009, η_p_^2^ = 0.19; MNI: 34, -20, 20; *F*_(1, 89)_ = 17.63, *P*_FWE_ = .025, η_p_^2^ = 0.17) showed an inverted pattern with increased responses to fast touch compared with slow touch.

For the model comparing responders and non-responders, we found main effects of speed in the left caudate nucleus (MNI: -18, 20, 12; *F*_(1, 49)_ = 19.14, *P*_FWE_ = .029, η_p_^2^ = 0.29) and the left (MNI: -36, 0, 12; *F*_(1, 49)_ = 21.78, *P*_FWE_ = .012, η_p_^2^ = 0.32) and right posterior insula (MNI: 36, -16, 22; *F*_(1, 49)_ = 27.13, *P*_FWE_ = .002, η_p_^2^ = 0.37). While the cluster in the left posterior insula exhibited increased response to slow touch compared with fast touch, the reverse pattern was evident in the clusters in the right posterior insula and the caudate nucleus.

In accord with our main findings, the baseline analysis of the contrast [controls > patients] revealed significant effect in the bilateral caudate nucleus (MNI: -12, 20, 10; *t*_(1, 89)_ = 3.81, *P*_FWE_ = .041, *d* = 0.80; MNI: 8, 16, 6; *t*_(1, 89)_ = 4.20, *P*_FWE_ = .013, *d* = 0.88). Additionally, we found two significant clusters in the right posterior insula (MNI: 38, -2, 16; *t*_(1, 89)_ = 3.90, *P*_FWE_ = .030, *d* = 0.82; MNI: 42, -8, 4; *t*_(1, 89)_ = 3.79, *P*_FWE_ = .042,*d* = 0.79). However, no significant effect was found for the nucleus accumbens or any of the other regions of interest. Baseline analysis did not reveal any significant effects for the contrast [patients > controls], nor for the comparison of responders and non-responders to antidepressant treatment ([responders > non-responders], [non-responders > responders]).

*Moderation effects*

We found that none of our predictors significantly moderated the effect of group or treatment response on any of our behavioral ratings (all *P*’s > .05). For the moderation analysis of the fMRI results, we found that childhood trauma questionnaire (CTQ) scores had a moderating influence on the effect of group on parameter estimates in the right nucleus accumbens (*t*_(89)_ = 2.17, *P =* .033). The Johnson-Neyman technique revealed that the relationship between group and parameter estimates in the right nucleus accumbens was significant when CTQ scores were less than 30.33. This suggests that the occurrence of clinical depression does not impact the response of the nucleus accumbens to social touch in people that have suffered from more severe childhood maltreatment. No significant moderation effects were observed for parameter estimates in any other region.

**Supplementary Tables**

**Table S1. Demographic and clinical data for responders and non-responders to treatment**

|  | Responders (n = 23) | Non-Responders (n = 30) | *P*-value |
| --- | --- | --- | --- |
| Sex (male/female) | 12/11 | 15/15 | 1.000 |
| Age (in years) | 43.57 (13.73) | 40.07 (12.60) | 0.340 |
| Education (in years) | 15.39 (3.87) | 16.27 (6.39) | 0.565 |
| Handedness (left/right) | 2/21 | 2/28 | 1.000 |
| Duration current depressive episode (in years) | 4.51 (4.94) | 4.77 (7.60) | 0.889 |
| Number of depressive episodes | 2.36 (2.17) (n = 21)^a^ | 3.79 (3.16) (n = 26)^a^ | 0.084 |
| **HDRS-17** |  |  |  |
| Baseline | 16.48 (5.33) | 17.87 (5.86) | 0.378 |
| After treatment | 5.61 (2.39) | 13.73 (5.09) | < 0.001 |
| Improvement (in percent) | 65.36 (12.14) | 21.26 (22.11) | < 0.001 |
| **BDI-II** |  |  |  |
| Baseline | 32.04 (9.52) | 34.33 (8.14) | 0.350 |
| After treatment | 13.35 (7.31) | 23.83 (10.91) | < 0.001 |
| Improvement (in percent) | 57.96 (18.82) | 29.23 (28.14) | < 0.001 |
| 4 weeks after treatment | 19.13 (12.08) | 24.96 (10.64)  (n = 27)^a^ | 0.076 |
| 8 weeks after treatment | 20.73 (10.43) (n = 22)^a^ | 26.19 (10.65)  (n = 27)^a^ | 0.078 |
| 12 weeks after treatment | 23.77 (8.42) (n = 22)^a^ | 24.92 (11.36)  (n = 24)^a^ | 0.702 |
| CTQ baseline | 42.57 (13.74) | 47.00 (17.95) | 0.330 |

Values are given as frequencies or as means (SD). The *P*-values report the significance levels reached for independent *t*-tests or Fisher’s exact tests comparing groups or for paired *t*-tests comparing improvement within patients. ^a^ Sample size in parentheses indicates number of complete responses. The significance threshold was set at *P <* .05.

**Table S2. Whole-brain activation in healthy controls (Touch vs. No Touch)**

| Region | Right/left | | Cluster size (voxels) | *t*-score | MNI Coordinates | | | *P*-value |
| --- | --- | --- | --- | --- | --- | --- | --- | --- |
|  |  |  |  |  | x | y | z |  |
| **Touch > No Touch** | |  |  |  |  |  |  |  |
| Insula | | L | 14977 | 15.30 | -40 | -4 | 8 | < 0.001 |
| Postcentral Gyrus | | L |  | 14.18 | -64 | -22 | 22 | < 0.001 |
| Supramarginal Gyrus | | L |  | 13.72 | -56 | -22 | 18 | < 0.001 |
| Supramarginal Gyrus | | R | 9303 | 13.56 | 52 | -28 | 24 | < 0.001 |
| Supramarginal Gyrus | | R |  | 12.70 | 60 | -24 | 22 | < 0.001 |
| Postcentral Gyrus | | R |  | 12.61 | 18 | -42 | 74 | < 0.001 |
| Middle Temporal Gyrus | | R | 842 | 11.75 | 54 | -60 | 4 | < 0.001 |
| Cerebellum VI | | R | 1669 | 10.53 | 24 | -52 | -22 | < 0.001 |
| Cerebellum VI | | R |  | 8.78 | 18 | -72 | -18 | < 0.001 |
| Cerebellum VI | | R |  | 8.53 | 24 | -64 | -20 | < 0.001 |
| Middle Temporal Gyrus | | L | 609 | 9.15 | -50 | -66 | 6 | < 0.001 |
| Cerebellum VI | | L | 167 | 9.09 | -24 | -62 | -22 | < 0.001 |
| Cerebellum VI | | L |  | 8.84 | -16 | -70 | -20 | < 0.001 |
| Thalamus | | L | 309 | 7.34 | -12 | -16 | 4 | 0.001 |
| Middle Frontal Gyrus | | R | 466 | 7.02 | 44 | 48 | 8 | 0.002 |
| **No Touch > Touch** | |  |  |  |  |  |  |  |
| Inferior Parietal Gyrus | | L | 906 | 7.52 | -36 | -76 | 42 | < 0.001 |
| Angular Gyrus | | L |  | 7.33 | -36 | -66 | 38 | 0.001 |
| Precuneus | | R | 1131 | 6.69 | 8 | -48 | 40 | 0.005 |
| Middle Cingulate Cortex | | L |  | 6.26 | -2 | -42 | 44 | 0.018 |
| Middle Temporal Gyrus | | L | 629 | 6.39 | -52 | -38 | -2 | 0.012 |
| Middle Temporal Gyrus | | L |  | 6.33 | -62 | -42 | -4 | 0.014 |
| Inferior Occipital Gyrus | | L | 157 | 5.91 | -22 | -92 | -6 | 0.045 |

An initial cluster-forming height threshold of *P* < 0.001 was used. Only cluster with FWE-corrected *P*s < 0.05 on peak level are listed. Abbreviations: MNI, Montreal Neurological Institute
